# Implementation of the Bristol Community Approach to improve vaccine confidence – A realist evaluation

**DOI:** 10.64898/2026.11.26.26367909

**Authors:** Megan N Pardoe, Taru Silvonen, Christalla Pithara-McKeown, Geoff Wong, Robyn Heath, Clare E French, Matthew Hickman, Lucy Yardley, Alexandra L. Creavin, Sarah Denford

**Author notes:** Corresponding author: Taru Silvonen. joint first authors (Megan N Pardoe, Taru Silvonen). joint senior authors (Alexandra L. Creavin, Sarah Denford).

## Abstract

**Background:** Measles, Mumps and Rubella (MMR) vaccination rates are lower than required to provide community protection both in England and in the study location Bristol, North Somerset and South Gloucestershire (BNSSG). To tackle this, the BNSSG Integrated Care System (ICS), in partnership with local community organisations, applied an asset-based approach to vaccination. This led to the development of the Bristol Community Approach (BCA), which builds on learning from the delivery of the COVID-19 Mass Vaccination Campaign in Southwest England.

**Methods:** This realist evaluation applies a case study approach to understand what works for whom and in what context in the delivery of the BCA focusing on MMR vaccination. We conducted 21 interviews with service providers involved in the delivery of the BCA, including 7 community organisations. Additionally, 46 public participants took part in 6 interviews and 5 focus groups. Our analysis led to the development of the programme theory in the form of context-mechanism-outcome-configurations.

**Results:** We identified six interconnected themes: 1) boundary spanners as trust brokers; 2) tailoring and delivering accessible engagement; 3) collaborative infrastructure and capacity; 4) threats to trust and engagement; 5) community norms and beliefs; 6) structural constraints on trust-building. The final program theory comprise eight ‘context-outcome-mechanism-configurations’ that span the six themes.

**Conclusion:** Effective community-based approaches are required to address several specific needs simultaneously. Long-term investment in these approaches is required to sustain trusted relationships between underserved communities and healthcare institutions.

## 1. Introduction

Improving vaccination coverage for measles, mumps and rubella (MMR) is an urgent public health concern in England. Vaccination rates (83.6%-91.8% across vaccination groups in 2024/25 (1)) are below the 95% World Health Organization (WHO) recommendation (2). Furthermore, evidence suggests that childhood MMR coverage rates will continue to fall (3). Only 88.5% of 2-year-olds in Bristol, a city in South West England, received one dose of MMR in 2023/24; lower than both the national average (88.9%) and the South West average (92.5%) (4 figure 6).

Childhood MMR vaccination uptake is not uniform across the region: uptake in Bristol ranges from 82% - 92% (4) with some demographic groups experiencing particularly low coverage, including university students; Somali communities, global majority communities such as Chinese and Eastern European communities as well as people living in areas of high deprivation (5, 6).

During recent measles outbreaks in 2018 and 2024 multiple cases occurred in Bristol (139 of the 971 cases in England in 2018; 52 of 2,911 in 2024) (7, 8). To mitigate the risk of future outbreaks, supporting uptake of the MMR vaccination was a key priority.

### Bristol Community Approach

The Bristol Community Approach (BCA) is an asset-based approach developed over 15 years to improve vaccination uptake in underserved communities in Bristol, North Somerset and South Gloucestershire (BNSSG) (Figure1). The approach was strengthened during the COVID-19 pandemic, when the urgency of delivering vaccinations led to closer partnerships between healthcare professionals, public health teams, and community organisations, supporting effective and accessible vaccine delivery across the region (9).

**Figure 1.**
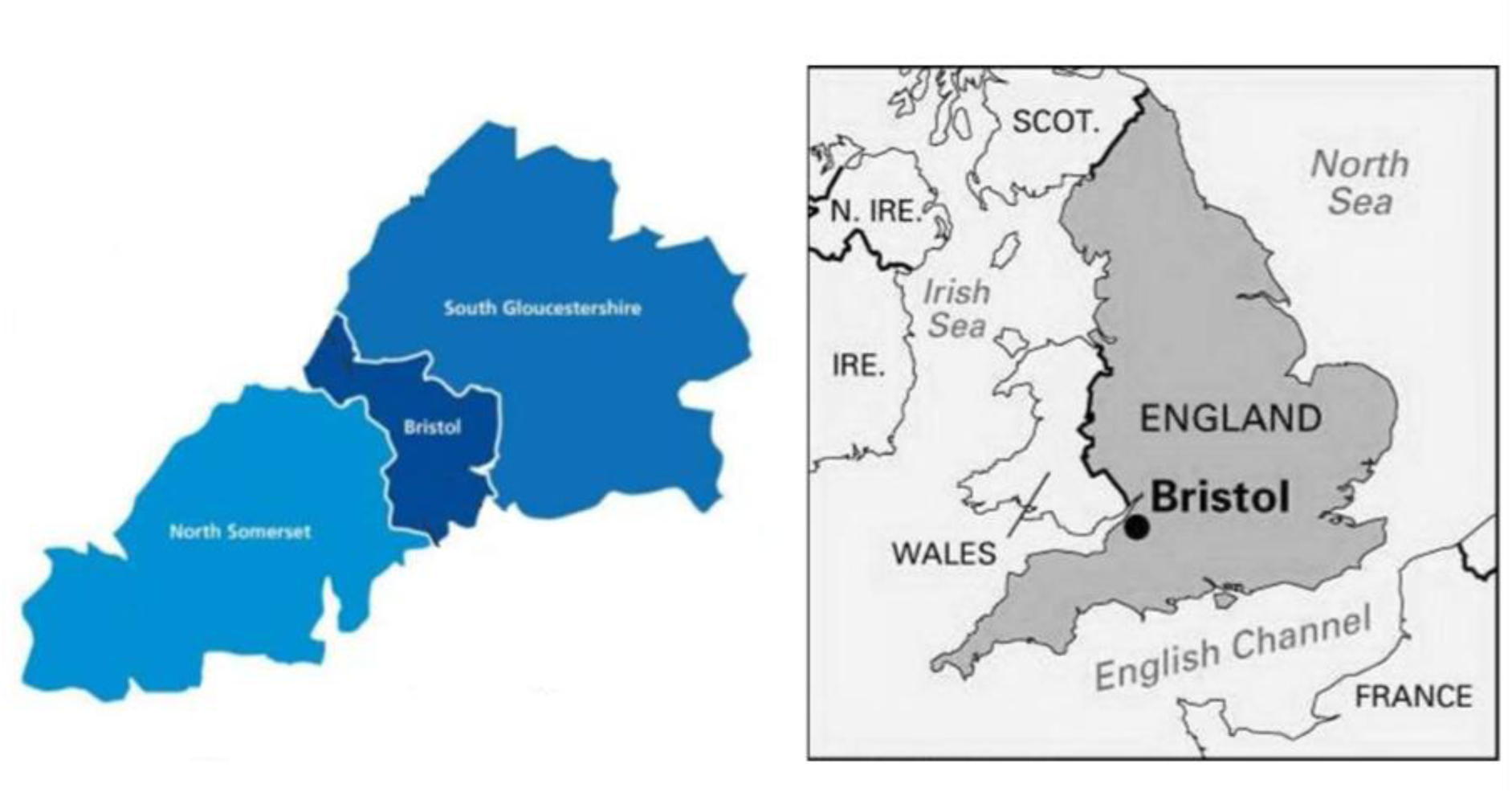
Map of Bristol, North Somerset and South Gloucestershire alongside a map of England

The BCA focuses on communities experiencing greatest inequalities and lowest uptake identified through local data; including people living in proportionally deprived areas; people from ethnic minority groups; people from Somali communities and students. The approach uses community engagement to develop culturally appropriate and accessible approaches to vaccine delivery. Building on success of this approach during the COVID-19 pandemic, the BCA has been further adapted to support MMR and, more recently, MMRV vaccination in underserved communities in BNSSG Integrated Care System in response to recent measles outbreaks (10). In brief, funding was delivered to nine voluntary organisations that have existing relationships with communities with low vaccine uptake. Organisations were invited to use the funding to support their communities to make informed decisions about whether or not to have the vaccine. An initial programme theory (IPT) (see figure 2), was developed through discussion with methodological and subject experts, and through collating information from 26 documents shared by BCA delivery team and community organisations involved in BCA activities. A data request form was also sent to the VCSEs involved in BCA delivery to seek further information on the activities VCSEs delivered.

**Figure 2.**
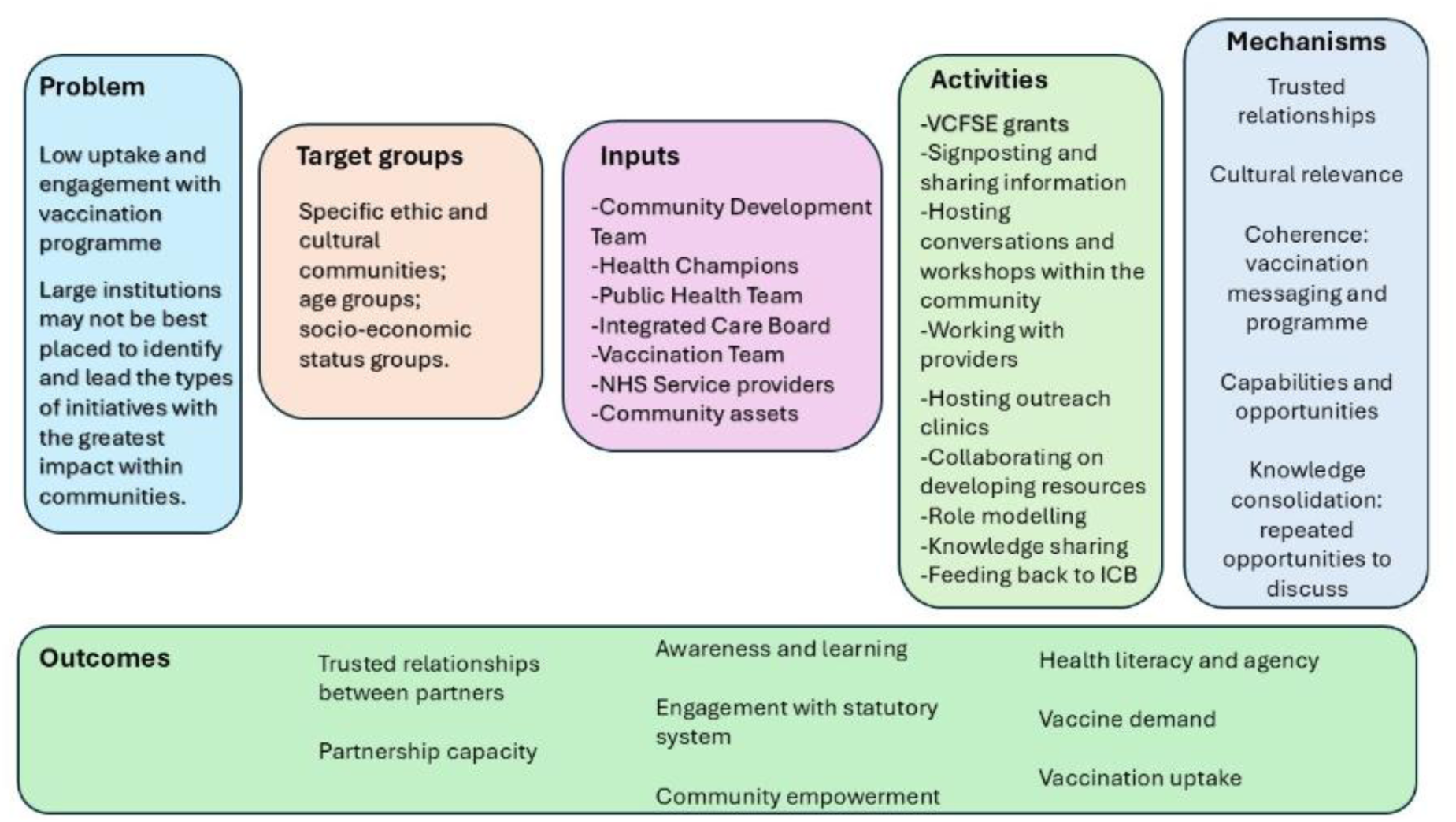
Bristol Community Approach - Initial Programme Theory

### Aims and objectives

The project’s overarching aim was to understand in what contexts, for whom and why the BCA can lead to change in vaccine attitudes, confidence and uptake in underserved communities. It reports on a realist evaluation of the BCA drawing on qualitative data from service providers (SPs) and public participants (PPs).

This evaluation contributes to a growing knowledge-base which is particularly relevant to healthcare commissioners, service providers, public health experts, researchers and consultants. While there is indicative evidence of the impact of local and tailored community approaches in public health (11–13), at time of writing there are no published detailed evaluations of asset-based and co-delivered vaccination initiatives in the UK that focus on more than one target population group.

## 2. Methods

This paper reports one component of a focussed realist evaluation (RE) study, namely, development of the programme theory (IPT) and its constituent ‘context-mechanism-outcome’ configurations (CMOcs) (14). RE allows exploring the circumstances in which and how an initiative works for different actors (15, 16). As the BCA involves several different service providers and socio-demographically differing communities as service users, applying an RE approach enables focusing on the differing contexts in which these actors are engaging with the BCA. Subsequent papers reporting a mixed methods study and an economic evaluation are presented elsewhere (see 17).

### 2.1 Participant recruitment

#### Setting

This research was conducted in two geographical locations where BCA was delivered: Bristol and Weston-super-Mare. These areas cover 1 million residents, of which some 16% are part of global majority ethnic groups (18).

#### Recruitment of Service providers

Service providers were eligible to take part if they worked at one of the nine identified organisations that received funding to support uptake of the MMR. A member of the study team initially approached the organisations involved in the BCA via email to invite individuals to take part in an interview. Those interested in taking part were sent a copy of the information sheet and were invited to identify a time for an online interview.

#### Recruitment of Public participants

Public participants from communities with low vaccine uptake were recruited via recruitment flyers that were shared via BCA community organisations. Recruitment materials were shared mostly digitally through WhatsApp groups and via email but also by handing out printed flyers.

Additional recruitment was conducted through existing contacts with local organisations and community-based groups, mailing lists, noticeboards, and in-person. Printed flyers were also provided to local venues with high footfall such as supermarkets and libraries.

Potential participants registered their interest using an online form, sharing their availability and preferences for mode of participation (online or in person; one-to-one interview or focus group). This resulted in 6 participants taking part in online interviews and 40 participants joining 5 focus groups (FGs).

### 2.2 Data collection

#### Service provider interviews

Data collection took place between September 2025 and August 2026. We conducted 21 interviews with service providers (SPs); 9 with VCFSEs staff and 12 with clinicians, health care professionals (HCPs) and public health specialists from local BNSSG area and local authorities. All interviews were carried out online using MS Teams. Interviews lasted approximately one hour and followed a semi-structure topic guide. All participants were offered a £25 voucher as compensation for their time.

#### Public participant interviews and focus groups

Data collection took place between December 2025 and March 2026. All interviews (N=6) were held online using MS Teams. FGs were conducted either online (N=2) or in person (N=3). All participants (6 interviewed, 40 attended FGs) were given either a print or digital copy of the participant information sheet. Those who were contacted digitally were asked to complete a digital consent form; we also provided printed forms so that all participants could provide written informed consent prior to taking part.

FGs duration was approximately 1 hour and were facilitated by two to three members of the research team. Interviews were shorter at 30 minutes and conducted by one of the researchers. Both FGs and interviews explored participants’ knowledge and awareness of MMR vaccine. The sessions started with open-ended questions and focused also on prior information of both vaccines, in addition to exposure, concerns, and motivations relating to vaccination decisions. All participants received a £25 voucher as a thank you for their time.

## 3. Data analysis

All interviews were transcribed using the automated transcription in MS Teams, cleaned, anonymised and reviewed prior to analysis.

Data were then analysed using codebook thematic analysis. Initially, a coding framework was created based on the IPT, preliminary reading of reading interview transcripts and discussion with the study team. Data were then categorised against the coding framework using Microsoft Excel. Codes were then reviewed and similar codes grouped, named and defined. The final stage of the analysis focused on creating ‘if-then’ statements The IPT was refined through a comparison of the CMOcs against the IPT to elucidate the interventional strategies that were central to the implementation of the BCA.

### Patient and public involvement and engagement

Thirteen public contributors were recruited through community organisation and the People In Health West of England and supported the conception and design of the study through discussions in an online meeting with the study team in February 2026.

### Ethical approval

Ethical review was obtained from the University of Bristol’s Research Ethics Committee (reference number: 25856).

## 4. Results

We identified eight overarching ‘context-outcome-mechanisms-configurations’ (CMOcs) which explained key aspects of the BCA.

### 4.1 Building and sustaining trust: Boundary spanners as trust brokers

As BCA builds on work initiated during the COVID-19 pandemic, the continuing trust-building leverages previously formed relationships and acts as the foundation underpinning the BCA. Interviews with both SPs and PPs highlighted that these trusting relationships enable sharing vaccination messaging with underserved communities, helping to overcome deeply rooted lack of trust in local and national institutions.

In BCA, boundary spanners are healthcare and local authority professionals such as community champions, community leaders, faith leaders and healthcare professionals embedded in communities who through continued engagement with their local communities can build an understanding of local concerns and needs. Their experiences, expertise and position as community insiders, who are respected and trusted by the community, enable them to bridge the gap between healthcare professionals or healthcare institutions and the service user communities. Boundary spanners thus provide an intervention strategy through which trust can be operationalised to support engaging with different communities in ways that support forming a better understanding of vaccine information.

#### Configuration 1

When boundary spanners help disparate healthcare organisations and community groups to build working relationships with each other (C) collaboration between these organisations facilitates effective sharing of vaccination information (O) because the organisations and groups develop confidence in the intentions and abilities of each other (M).

As a ‘boundary spanner’ from a community organisation working especially with Somali communities explained, connecting the NHS team with organisations that work with different communities brought a diverse approach to sharing vaccine information.

The collaborations with VCSE organisations highlighted the additional needs within different communities and how organisations that were directly connected to the communities they serve were able to bridge any gaps during vaccination events. The provision of specialist translation supported making key messages applicable to the context of the attendees of a vaccine talk, lowering the barrier to get vaccinated within the Chinese community.

Referring to their experience at a community event that had a local vaccine champion present, a local authority employee explained how the way the community organisation delivered vaccination messaging differed from the approach they were used to. Seeing the reaction of the community members and the effectiveness of what seemed an unusual approach instilled confidence in them about the importance of community functions.

Boundary spanners act as messengers, translators, connectors, and advocates, representing their local communities. While boundary spanners can help share messages with the communities they represent, they can also introduce new ways of working to the organisations they collaborate with as they know the needs of the community. This can include translation to make the information understandable by non-English speakers but also activities that soften the delivery of the message by making vaccination a part of other community activities, giving vaccine events, held in the community, familiarity and a sense of belonging to the local community. See table 1 for data relating to this configuration.

**Table 1.** Verbatim quotes of data examples for configuration 1.

|  |  |
|---|---|
| Quote 1 | *“There was other community organisations working promoting the MMR vaccinations, for example Chinese organisations and Asian-led organisations, so the NHS were working with them, with organisations led by communities and there was a diversity inside diversity” (P05).* |
| Quote 2 | *“I think one of the most important is interpretation and then guiding people like if we have a vaccine talk and then we'll have the drop in vaccine session in another room so they will be guiding people to the room. The NHS, they didn't bring any interpreter, so we need to interpret, otherwise they can't have their jab” (P15).* |
| Quote 3 | *“When we went there [community event]... people were dancing and music was playing and then people are queuing for the vaccine...it just showed that sometimes the system needs to take that community leadership and trust the communities because the community can do better” (P20).* |

### 4.2 Collaborative Infrastructure and Capacity

Partnerships between healthcare providers and voluntary, community and social enterprise (VCSEs) organisations contributed to the creation of collaborative infrastructure. This infrastructure allowed learning to be shared between the different organisations, enabling the delivery of appropriate vaccination events and activities.

#### Configuration 2

When NHS, local authority and VCSE organisations facing resource limitations are able to share experiences, advice and materials to support each other build connections (C), community approaches are more likely to be delivered effectively (O) because of greater resilience (M).

Interviews with SPs showed that organisations were able to support sharing vaccine information by learning from each other but also sharing materials and advice about how to engage with specific communities. The local authority-based communities’ team was one key actor enabling further work with communities through channels that were known to be effective.

One of the VCSE organisations involved in delivering outreach activities especially within inner city Somali communities felt that the collaborations between healthcare organisations, VCSEs and the community led to mutual benefit. Relationships between organisations deepened while communities benefitted from increased engagement with HCPs.

All SPs recognised funding limitations and the challenges these brought to their work. Notably, SPs felt that building relationships both with organisations and the community takes time, whereas funding tended to be available for short-term and ‘ad hoc’. Resource was acknowledged to be sparse, which meant that further close collaboration was needed to ensure the work could be continued. See table 2 for data relating to this configuration.

**Table 2.**
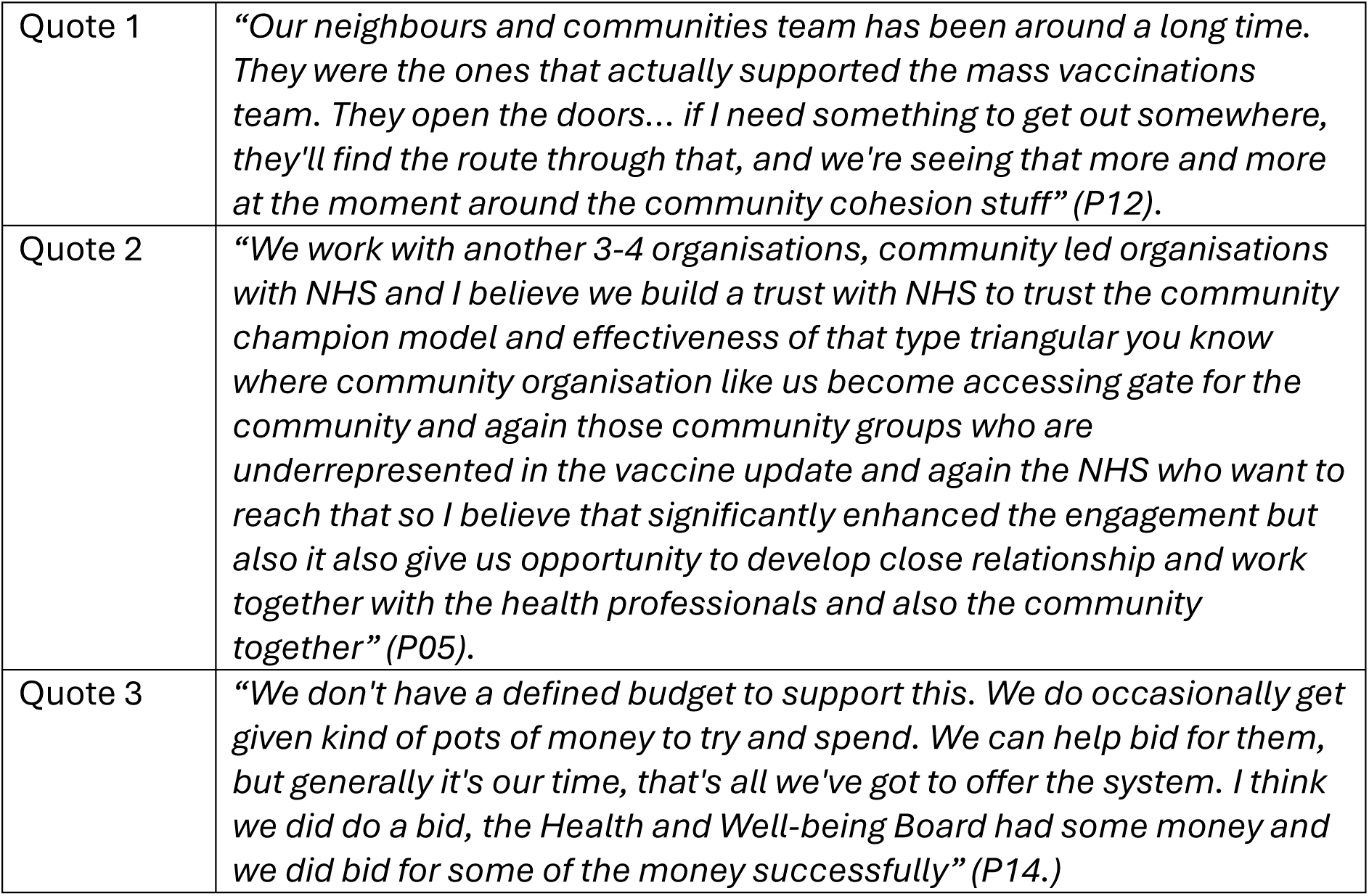
Verbatim quotes of data examples for configuration 2.

|  |  |
|---|---|
| Quote 1 | *“Our neighbours and communities team has been around a long time. They were the ones that actually supported the mass vaccinations team. They open the doors... if I need something to get out somewhere, they'll find the route through that, and we're seeing that more and more at the moment around the community cohesion stuff” (P12)*. |
| Quote 2 | *“We work with another 3-4 organisations, community led organisations with NHS and I believe we build a trust with NHS to trust the community champion model and effectiveness of that type triangular you know where community organisation like us become accessing gate for the community and again those community groups who are underrepresented in the vaccine update and again the NHS who want to reach that so I believe that significantly enhanced the engagement but also it also give us opportunity to develop close relationship and work together with the health professionals and also the community together” (P05)*. |
| Quote 3 | *“We don't have a defined budget to support this. We do occasionally get given kind of pots of money to try and spend. We can help bid for them, but generally it's our time, that's all we've got to offer the system. I think we did do a bid, the Health and Well-being Board had some money and we did bid for some of the money successfully” (P14.)* |

### 4.3 Tailoring and delivering accessible engagement

Community organisations working in partnership with other providers such as local authority public health teams, HCPs and community organisations can provide the community with knowledge needed to make informed decisions about vaccination. The community organisations themselves may be trusted by their local communities, but the communities may lack trust in institutional providers. It is important that these relationships are built and developed to gain the trust of the targeted community groups. Once community organisations facilitate relationship building between communities and healthcare providers, trust can start to shape, as organisations can adapt engagement approaches to local needs.

#### Configuration 3

When boundary spanners share information with BCA communities that is adapted to formats and languages, they find accessible and provide opportunities to discuss the information with people (C), people are more likely to act upon the vaccine information (O) because they can understand it (M).

Boundary spanners are key in helping to share accessible knowledge amongst communities, where something else is needed in addition to the information and resources that exist as standard NHS care.

One community member felt that attending MMR sessions organised by their community organisation allowed them to find out the information they needed. This led to feeling they could make an informed decision about MMR vaccination uptake for themselves and their children.

A provider highlighted that using statistics and speaking from a higher position of knowledge does not help reduce people’s fears especially with the amount of information that circulates on social media. They felt that there is a vast amount of information which can exacerbate fears as social media is so readily accessible by many people.

In addition to ensuring that messaging is in the most accessible format, BCA shows that taking both vaccination opportunities and activities where vaccines can be discussed openly to the local settings is valued by the communities and has a greater impact than statistical information. See table 3 for data relating to this configuration.

**Table 3.** Verbatim quotes of data examples for configuration 3.

|  |  |
|---|---|
| Quote 1 | *“What's the right thing for 80% of the population and then what you need to do for that 20% that really need it? And so you've got to think about things on a really, really, really small local level, but also kind of that scale needs to needs to work up as well, so it's trying to cover all your bases” (P16)*. |
| Quote 2 | *“When we did the talk last time, the group session, I ended up having the MMR vaccine myself and gave it to the rest of my children that were missing one lot of vaccination. So that did really help me to make an informed decision. So these sessions are helpful to be honest.” (FG5)* |
| Quote 3 | “Social media processes that exacerbate fears...I can kind of speak humanly about frustrations with those alongside. So I think these things are really emotive, I think if you just answer in statistical terms and try and speak from some higher position of knowledge and certainty, then people don't really feel met” (P13). |

#### Configuration 4

When vaccine talks and drop-in clinics are held in spaces within the community which are familiar, trusted and easily accessible (C) people may be more likely to attend (O) because they feel comfortable to do so (M).

One community organisation saw the value in giving people the space and opportunity to ask questions from HCPs in a space that felt trusted. They felt that people may not ask questions if they had gone to their GP, where the environment is less familiar and they are not surrounded by relatable people.

Community organisations believed that their community trusted in them and what they were offering which allowed good turnout for drop-in vaccination sessions and talks. They held the majority of sessions in the same space that was easily accessible by the community.

One public participant shared their experience about receiving the flu vaccine yearly at their local mosque. This was a convenient, accessible and familiar location in which nurses also offered a gelatine-free flu vaccine, meeting the needs of the community.

Organising vaccine talks or offering vaccination in accessible, familiar spaces that local communities perceive to be safe places can help reduce several vaccination access barriers simultaneously. See table 4 for data relating to this configuration.

**Table 4.** Verbatim quotes of data examples for configuration 4.

|  |  |
|---|---|
| Quote 1 | *"This is why it's important and give people the space to ask questions. I think because they're in a space that they're surrounded by people, that they feel maybe are a bit more like them and in a space that is trusted, they're able to ask questions. But perhaps they wouldn't if they were in their own personal primary care setting" (P04).* |
| Quote 2 | *"I think one of the thing is the trust that they have with our community. So what we offered, they would think that's a safe choice and then before the drop-in session they give a talk. So they also learn about the knowledge. Some of them maybe they are still not sure about ‘should I have a vaccine or’ but that gives them an opportunity to talk to people and talk to the health profession and ask, ‘oh, should I have a vaccine’ which you don't normally get people to talk to you about " (P15).* |
| Quote 3 | *“I think it was the flu jab, the one where it has gelatine in it. So they had the halal version of it, and the nurses came to the mosque to do it...I think if I had had to take them elsewhere, or [go] to a location that is not familiar, or something like that, I probably wouldn't have done it. I would have just been like, that's got gelatine, and then we won't have that. But because it was like a pop-up service, the mosque is like a 5-minute walk away, and it was also convenient. We were in and out within like 10 minutes. So we've been doing that every year because they do it every year” (PP4).* |

### 4.4 Threats to Trust and Engagement

Historical and social drivers of mistrust include factors that undermine confidence in vaccination programmes and healthcare systems before any engagement begins. The data show that BCA communities, apart from the student population, are characterised by a general mistrust of the NHS, some of which is fuelled by previous negative experiences as well as legacies of being excluded from healthcare.

#### Configuration 5

When HCPs working with trusted community organisations provide local opportunities to discuss communities’ specific vaccine concerns (C) people are more likely to consider the information they are provided with (O) because the HCPs are better placed to overcome any historical mistrust (M).

A SP from a VCSE organisation working with young people and Somali communities highlighted the lack of reassurance and opportunities to openly discuss vaccination related concerns as a barrier to building confidence in vaccination.

Community champions were found to be an effective way to provide trusting settings for discussing vaccines. This was especially the case with VCSEs working with global majority communities such as the Chinese and Somali community.

A SP from a VCSE working with deprived areas in inner city Bristol noted that bringing vaccination discussions to local settings addresses several barriers simultaneously. The importance of being able to have a conversation in an unpressured setting with someone people can relate to was echoed by several of the SPs as well as PPs from different global majority groups. See table 5 for data relating to this configuration.

**Table 5.**
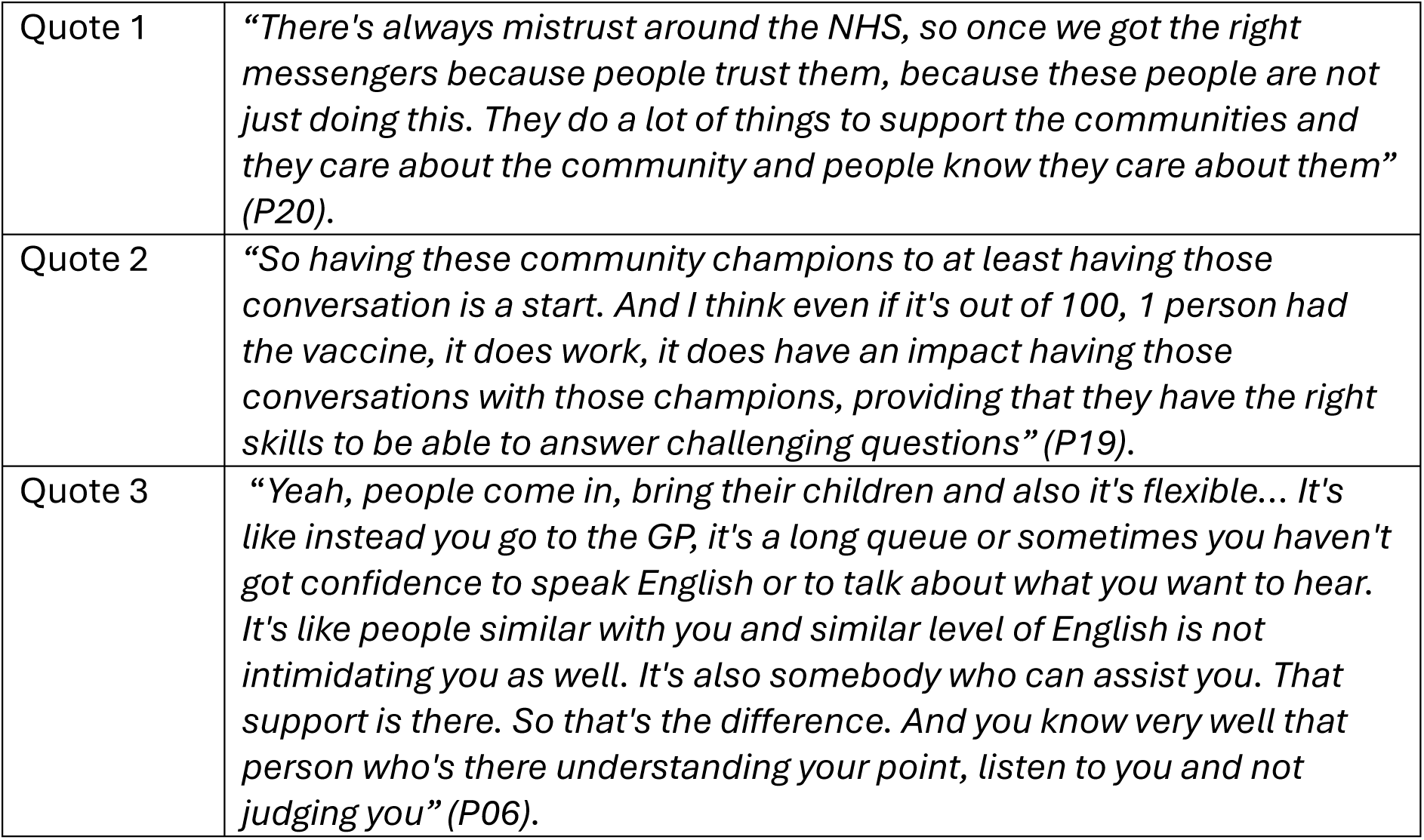
Verbatim quotes of data examples for configuration 5.

|  |  |
|---|---|
| Quote 1 | *“There's always mistrust around the NHS, so once we got the right messengers because people trust them, because these people are not just doing this. They do a lot of things to support the communities and they care about the community and people know they care about them” (P20).* |
| Quote 2 | *“So having these community champions to at least having those conversation is a start. And I think even if it's out of 100, 1 person had the vaccine, it does work, it does have an impact having those conversations with those champions, providing that they have the right skills to be able to answer challenging questions” (P19).* |
| Quote 3 | *“Yeah, people come in, bring their children and also it's flexible... It's like instead you go to the GP, it's a long queue or sometimes you haven't got confidence to speak English or to talk about what you want to hear. It's like people similar with you and similar level of English is not intimidating you as well. It's also somebody who can assist you. That support is there. So that's the difference. And you know very well that person who's there understanding your point, listen to you and not judging you” (P06).* |

### 4.5 Community norms and beliefs

Both the SPs and PPs highlighted that the presence of several competing narratives made it difficult to identify trustworthy and accurate information about vaccines. Alternative explanations and information sources that challenge the official messaging about vaccines included especially rumours in the community about the link between MMR and autism. This contributed to the persistent negative perception of the vaccine which was often exacerbated by misinformation on social media.

#### Configuration 6

When people with concerns about the safety and potential harms of vaccines can discuss their concerns (C) they are more willing to consider vaccination uptake (O) because they are more likely to feel reassured about the safety of the vaccination (M).

Especially the Somali communities had concerns about the link of MMR vaccinations to autism, which was thought to be because of the increasing presence of autism amongst the Somali population.

The data indicate that concerns are also common in other global majority communities. Addressing the plurality of competing narratives requires consistent sharing of vaccination information in settings where PPs can make an informed decision to get vaccinated once they feel ready to do so. A VCSE organisation supporting Chinese communities highlighted the positive impact holding vaccination talks could have.

The data suggest that the supportive process in terms of addressing safety concerns across different communities is sharing relevant information by trusted individuals who can support people make their own decisions on vaccination uptake. See table 6 for data relating to this configuration.

**Table 6.** Verbatim quotes of data examples for configuration 6.

|  |  |
|---|---|
| Quote 1 | *“you know, [there is an] increasing number of autism in Somali community, which is more known than before. A lot of people, they got doubts. Some of them, they read articles or research, you know, research about MMR vaccinations and it is linked with autism... what I have seen, majority of the discussions were relating to MMR vaccination is on autism and a lot of people didn't know, a lot of people that haven't got that reassurance to take the vaccine” (P05).* |
| Quote 2 | *“I don't think that a lot of people register with the vaccination when we promote it, and then I think they're quite unsure but they are willing to come for the talk and after they listen to the talk and then they come forward and take the vaccination” (P15).* |
| Quote 3 | *“When we talk to people, there were people who felt more confident in allowing their children take the vaccine. I know we did a lot of, we did a lot of phone calls for the GPs and you know, booked in people to come in for and have the MMR vaccine” (P19).* |

### 4.6 Structural Constraints on Trust-Building

#### System-level factors

Interviews with SPs highlighted system-level factors that make it difficult to establish and maintain the trusted relationships that are pivotal for the delivery of BCA. The short-term availability of funding and how this would further limit staff capacity was felt to undermine the ongoing work required to build and maintain trust.

#### Configuration 7

When organisations have limited capacity and short-term funding (C) maintaining collaborative relationships between HCPs and community organisations becomes challenging (O) because they are limited in how far they can plan ahead (M).

From a delivery perspective, funding restrictions led to community organisations often being limited in what they can focus on in their work, even if this meant not following the routes to outreach that were considered to potentially be effective.

Similar pressures were faced in healthcare organisations as collaborations with community organisations and improvement of outreach work were additional to normal workloads. This meant that while staff were interested in focusing on outreach work, this could not be prioritised.

The mismatch between the time it takes to build relationships that through continuity support forming trust and the availability of short-term funding was felt to hinder forward planning, standing in the way of effective and sustainable outreach. See table 7 for data relating to this configuration.

**Table 7.** Verbatim quotes of data examples for configuration 7.

|  |  |
|---|---|
| Quote 1 | *“It took some time to develop those relationships. I think there’s two things: one because of our core purpose, which is [local church], and because of the kind of resource allocated to it, it meant that we couldn't. You know our purpose for developing a really strong relationship with, I can't even remember the name of the church now, but the different churches kind of that [we] were recommended to go out to, there wasn't that incentive to do that” (P03)*. |
| Quote 2 | *“I went out to the team and there were four people came forward with that interest...They have their normal workload, but it's just something they pick up when they have a quiet hour or time...and try and spread the word about vaccines and network with different community members and try and get their face known and trusted within communities” (P09)*. |
| Quote 3 | *“It doesn't work to panic rather than planning. You have to plan. It needs longer term thinking than actually saying we have £20,000, so partners in the community come and help us in that way. It's unsustainable and doesn’t lead to long term change... in order to myth bust those taboo subjects, you need far more resource and rooted in the community for many years in order to see difference” (P01)*. |

### Access and Engagement barriers

Both SPs and PPs highlighted practical barriers that limit vaccine uptake even when vaccination messaging is being delivered through boundary spanners in trusted settings. These barriers include difficulties accessing vaccines, booking appointments with HCPs as well as language barriers and general health literacy. These barriers hinder the delivery of community-based approaches and if not addressed can undermine the efforts to support vaccination uptake.

#### Configuration 8

When organisations understand, tailor and address different relevant barriers to getting vaccinated in community settings simultaneously (C) people are more likely to consider vaccination (O) because people can more effortlessly get vaccinated (M).

VCSEs working with global majority communities such as the Somali communities highlighted several simultaneous aspects that act as barriers to getting vaccinated. While translated leaflets may be effective with some people, this would not be the case for people with limited literacy. Furthermore, difficulty making appointments with HCPs can still undermine an intention to get vaccinated that has resulted from additional information. Understanding the specific barriers that communities face requires addressing multiple barriers simultaneously.

Because it can be difficult for organisations to pre-empt specific uptake barriers experienced by the communities they support, in-person conversations can help provide tailored support on a more case-by-case basis.

Identifying ways to address multiple barriers such as either combining vaccination talks and opportunities to ask HCPs questions with the option of subsequently getting vaccinated locally was felt by SPs to be an effective way to deliver BCA across different communities. See table 8 for data relating to this configuration.

**Table 8.**
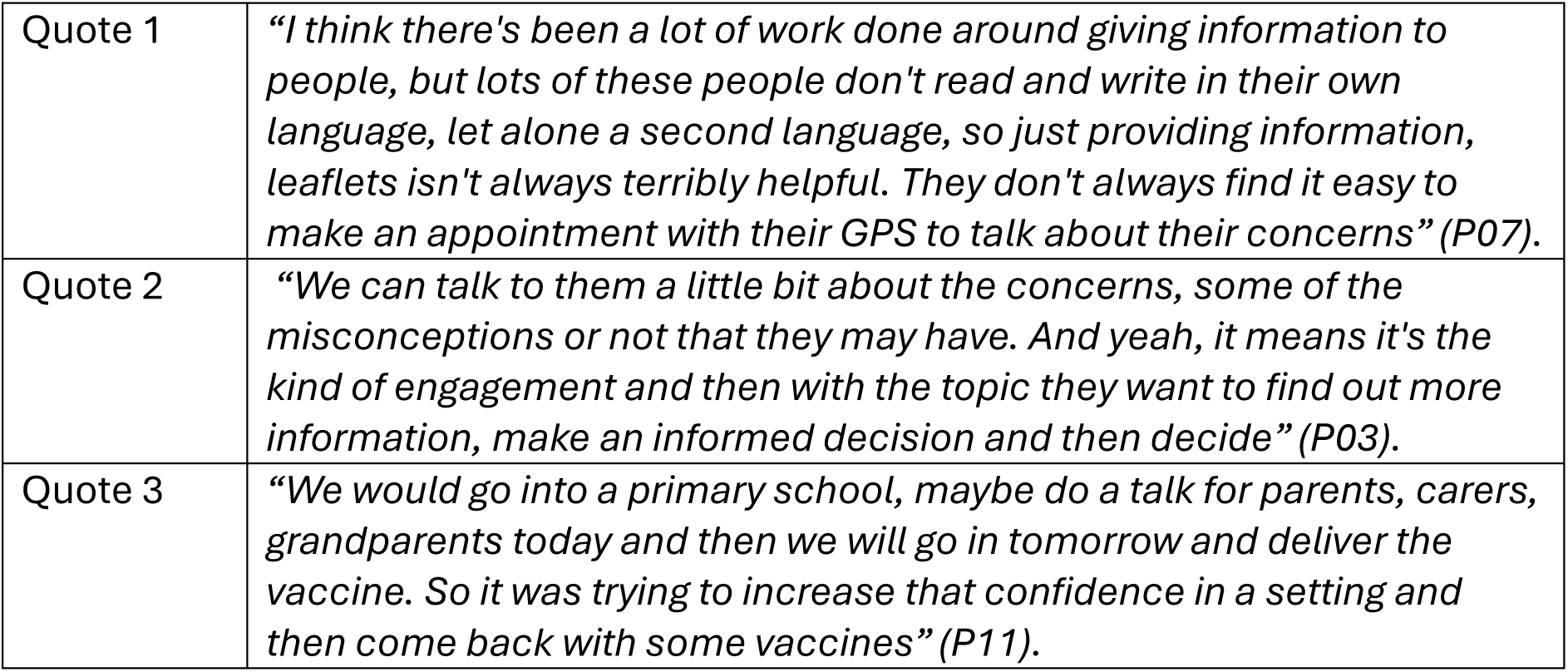
Verbatim quotes of data examples for configuration 8.

|  |  |
|---|---|
| Quote 1 | *“I think there's been a lot of work done around giving information to people, but lots of these people don't read and write in their own language, let alone a second language, so just providing information, leaflets isn't always terribly helpful. They don't always find it easy to make an appointment with their GPS to talk about their concerns” (P07).* |
| Quote 2 | *“We can talk to them a little bit about the concerns, some of the misconceptions or not that they may have. And yeah, it means it's the kind of engagement and then with the topic they want to find out more information, make an informed decision and then decide” (P03).* |
| Quote 3 | *“We would go into a primary school, maybe do a talk for parents, carers, grandparents today and then we will go in tomorrow and deliver the vaccine. So it was trying to increase that confidence in a setting and then come back with some vaccines” (P11).* |

## 5. Refined programme theory

Overall, the data suggest that sharing vaccine information through collaborations between healthcare professionals and local VCSE organisations has supported building trusting relationships between BCA organisations and the underserved communities in BNSSG. The data show that combining the use of trust brokering boundary spanners (configuration 1) with tailoring vaccination messaging (configuration 3) in ways that was meeting the norms and beliefs of local communities (configuration 5) whilst delivering engagement in accessible ways in familiar community locations (configuration 3) is linked to positive outcomes such as further engagement with messaging and intention or uptake of vaccination. Furthermore, especially PPs reported that embedding collaborative infrastructure and capacity across healthcare organisations and VCSEs (configuration 4) whilst acknowledging threats to trust building (configuration 5); system-level constraints such as short-term funding (configuration 7) and barriers to access and engagement within local communities (configuration 8) supported the delivery of BCA.

## 6. Discussion

This realist evaluation of BCA suggests that addressing low vaccination uptake in underserved communities requires providing communities multiple opportunities to discuss concerns in familiar settings, with trusted individuals facilitating the sharing of vaccine messaging. For some, this may also help address language barriers and relatability to the messenger.

Our data highlight the importance of continuously bridging institutional and community-based service provision to facilitate the sharing of vaccination messaging. This was highlighted through two configurations. Firstly, collaboration between organisations leads to trusting relationships between partnering health and community organisations. Boundary spanning also appeared to facilitate the acceptance of official health messaging, as communities saw their trusted organisations or individuals collaborating with HCPs (configurations 1, 5). Secondly, concerns and competing narratives are highlighted by the data as a barrier to considering vaccination, and trusted individuals are better placed to address these concerns (configuration 8). Whilst previous research highlights the importance of individuals known well in the community to facilitate trust in vaccination (19, 20), through BCA, we are able to provide practical advice about how these individuals can facilitate trust.

The observable connections between key configurations highlight interconnections between aspects of BCA. For example, effective collaboration between boundary spanners and healthcare organisations, can result in several barriers experienced by communities being addressed simultaneously (configuration 8). Sharing of information in safe and familiar locations further enhances the sharing of messages (configuration 4). While previous research has suggested that community-oriented interventions can help address vaccine confidence among global majority communities in the UK (21), our research highlights the importance of a multifaceted, multitargeted approach to vaccination; supporting access to both trusted information and access to vaccination. A key tension identified through this evaluation is that trust building takes time, effort and integrated activities. However, funding is usually available only on a short-term basis. This prevents the development of sustained partnerships and provision of activities that BCA indicate are essential for informed decision-making (configuration 7). For example, VCSE organisations were unable to engage with the communities when they did not have a previous existing relationship with the community. In this instance, efforts were focused on building new relationships before any vaccination activities could take place. The time-intensive nature of robust community engagement is also recognised by previous research, which urges for resources to be made available based on longer terms (19).

### Strengths and limitations

A key strength of this study was the collaboration between the academic research team and our clinical partners, who were able to support recruitment of diverse participants through sharing study materials and facilitating relationships with key contacts. This partnership also provided access to information about intervention components that was used to form the IPT. In addition, the involvement of 13 members of underserved communities in the design of the study were essential in ensuring that the materials and approaches were culturally appropriate.

The main limitation of this study is that interviews were conducted 12+ months after the activities, which may have resulted in recalling details inaccurately, and may have meant that members of the public did not have direct experience of intervention activities. Furthermore, despite efforts, we were unable to include the perspectives of one group targeted BCA due to a lack of contacts in that area. Of interest, BCA activities in this area were also challenging to deliver for similar reasons. Developing relationships within this area will be essential for both future research and interventions.

### Future directions for the BCA

Further research is now needed to consider the role of boundary spanners more widely in other areas and for additional public health behaviours. Whilst the current study indicates potential in this role, this now needs to be further developed through experimental study designs.

## 7. Conclusion

This realist evaluation has identified the contexts in which the multi-actor BCA can lead to change in vaccine hesitancy in underserved communities, emphasising the interconnected nature of trusting relationships between organisations to facilitate sharing vaccine messaging. Whilst the delivery of such community-based approaches is not without its challenges, any reduction to community outreach budgets should be considered carefully in light of the findings of this evaluation. While it takes time to see the results of community-based approaches, their influence in vaccination messaging to further uptake is promising.

## Data Availability

All data produced in the present study are available upon reasonable request to the authors

## Acknowledgements

SD, CEF, AC, LY, and MH acknowledge support from the NIHR Health Protection Research Unit in Evaluation and Behavioural Science at University of Bristol (NIHR207385). The Health Protection Research Unit (HPRU) in Evaluation and Behavioural Science at University of Bristol is part of the National Institute for Health and Care Research (NIHR) and a partnership between University of Bristol and UK Health Security Agency (UKHSA), in collaboration with the MRC Biostatistics Unit at University of Cambridge and University of the West of England.

We are grateful for the advice of clinical staff Clare Cook, Julie Northcote, Anne Morris and the feedback received from our patient and public involvement and engagement group. We thank the participants for taking the time to take part in this research. We acknowledge the work of temporary staff Luyun Liang and Rachel O’Donnell in helping cleanse interview transcripts.

## CRediT authorship contribution statement

Funding acquisition: AC, SD, LY, GW, CEF. Supervision: AC, SD. Methodology: SD, AC, LY, CEF and GW. Data collection: MP, TS, CPM, and RH. Data analysis: MP, CPM, GW and TS with support from SD. Writing – original draft: MP and TS. Writing - review and editing: all authors.

## Declarations

This study/project is funded/supported by the National Institute for Health and Care Research (NIHR) Three Research Schools Prevention Research Programme (Grant Reference Number NIHR20400 – Prev). The views expressed are those of the author(s) and not necessarily those of the NIHR or the Department of Health and Social Care. The National Institute for Health and Care Research (NIHR) Three Research Schools is a partnership between the School for Public Health Research (SPHR), the School for Primary Care Research (SPCR) and the School for Social Care Research (SSCR).

AC led the project to produce the MMR film: Let’s talk about MMR. The other authors declare no conflict of interest. The authors have no financial relationships with any organisations that might have an interest in the submitted work in the previous three years; no other relationships or activities that could appear to have influenced the submitted work.

